# Blood transcriptomic signature associated with hypertension in females across two human cohorts

**DOI:** 10.64898/2026.09.25.26364057

**Authors:** Chaoran Yang, Jianan Lu, Tanya Pretorius, Leticia Camargo Tavares, Francine Marques

**Affiliations:** Department of Pharmacology, Cardiovascular Disease Program, Biomedicine Discovery Institute, Faculty of Medicine, Nursing and Health Sciences, Monash University, Clayton, Australia; Victorian Heart Institute, Monash University, Clayton, Australia; National Centre for Epidemiology and Population Health, Australian National University, Canberra, Australian Capital Territory, Australia; Baker Heart and Diabetes Institute, Melbourne, Australia

**Keywords:** Sex difference, ETS family, hypertension, reproductive ageing

## Abstract

**Background:** Hypertension prevalence rises sharply in females after menopause, exceeding that of age-matched males in later life, conferring greater relative cardiovascular disease risk in females than in males. However, the underlying mechanisms remain poorly understood.

**Method:** We studied multi-tissue RNA-sequencing data from 193 CVD-free participants aged >55 years in the GTEx database and extended the findings to 1,605 age-matched Framingham Heart Study participants.

**Results:** We identified a dominant female hypertension co-expression signature in blood, expressed at lower levels in females with uncontrolled high blood pressure than in males (sex-by-hypertension interaction) in both cohorts. Genes contributing to this signature were enriched for post-transcriptional modification and insulin signalling and regulated by ETS family transcription factors (TFs). Ten ETS TFs showed the same female-specific hypertension association and together explained the signature’s expression. Moreover, in females, but not males, lower blood signature scores were associated with a pro-inflammatory transcriptome in the artery and kidneys.

**Conclusions:** Collectively, we identified the ETS-transcriptomic axis as a potential mechanism explaining sex differences in human hypertension.

**Graphic Summary:** 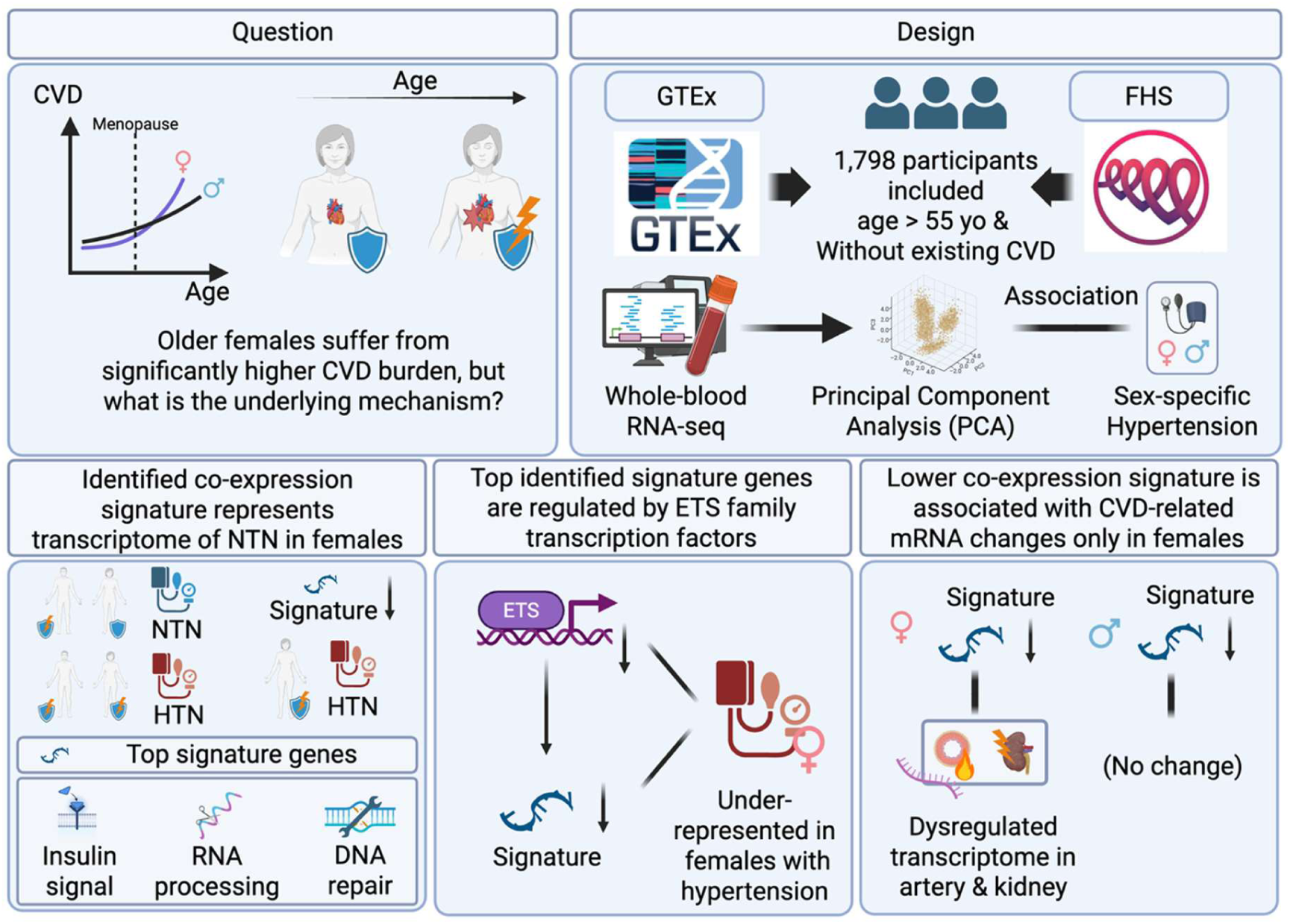

## Introduction

According to the Global Burden of Disease (GBD) study, cardiovascular disease (CVD) is the leading cause of death worldwide, with hypertension identified as one of the leading contributors.^1^ While premenopausal females exhibited a lower risk of hypertension and CVD than age-matched males, this protective effect progressively diminished with advancing age,^2,3^ especially following menopause.^4^ The menopausal transition is characterised by a substantial reduction of cardiovascular-protecting oestrogen levels.^5,6^ After the perimenopausal stage, blood pressure (BP) increases more rapidly in females than in age-matched males. The incidence of hypertension in postmenopausal females can even reach or surpass that of age-matched males.^7,8^ Moreover, CVD risk associated with hypertension is significantly higher in postmenopausal females compared to age-matched males and starts at a lower BP threshold in females.^9^ Despite these well-documented changes in hypertension and accompanied CVD risk across the female lifespan, cardiovascular risk in females has historically been underrecognised, contributing to disparities in healthcare access, underdiagnosis, and poorer clinical outcomes following CVD events.^10^

As the primary contributor to CVD, hypertension is a multi-factorial disorder. Ageing and menopause are key, non-modifiable risk factors.^7,8^ Systemic perturbations in gene expression in biological processes such as inflammation and fibrosis play an essential role in the pathophysiology of hypertension.^11^ Given the well-documented sex differences in hypertension and associated cardiovascular outcomes, molecular changes associated with hypertension may also differ between males and females. Indeed, a handful of studies have suggested that accelerated vascular ageing may contribute to the development of hypertension in postmenopausal women.^12,13^ Nevertheless, the molecular mechanisms underlying sex differences in hypertension remain poorly understood. This knowledge gap further constrains the development of effective female-specific strategies for the prevention and treatment of hypertension.

To address this gap, we analysed transcriptome data from two independent cohort studies, the Genotype-Tissue Expression (GTEx) and the Framingham Heart Study (FHS), to identify sex-specific transcriptional signatures associated with hypertension in the postmenopausal female population. We then analysed the biological pathways, genetic determinants, and upstream regulators underlying these signatures and associated systemic transcriptional changes to better understand sex-dependent mechanisms of hypertension.

## Methods

### Inclusion criteria

The Genotype-Tissue Expression (GTEx) is an NIH Common Fund-supported cohort study that includes transcriptome and epigenome data from up to 54 non-diseased tissue sites from body donation.^14^ GTEx participants aged over 55 years, widely considered representative of a population consisting predominantly of postmenopausal females by other studies,^15,16^ and age-matched males, were included. The hypertension diagnosis in GTEx was extracted from the previous medical record of participants by the GTEx project, defined as SBP exceeding 140 mmHg and/or DBP exceeding 90 mmHg.^17^ Participants without available whole blood RNA-seq data, clinical hypertension diagnosis records, records of alcohol consumption, smoking status, and diabetes diagnosis data, or with a history of cardiovascular diseases or current cancer diagnosis were excluded from the following analyses.

The FHS is a population-based prospective cohort focused on cardiovascular disease risk factors.^18^ In our study, we extracted FHS participants from the Generation II Exam 8 dataset with available whole-blood RNA expression data measured by the Affymetrix Human Exon 1.0 ST microarray. Of these participants, 1,646 were aged over 55 years and had no history of cardiovascular disease. Of these 1,646 participants, we excluded 6 without complete information from medical examination and interview and 17 premenopausal female participants. We also excluded 18 participants due to incomplete blood pressure (BP), discontinued BP-lowering medication, alcohol consumption, smoking status, or diabetes data, leaving 1,605 participants for the following analyses. Included FHS participants were then grouped by BP using the standard as follows: uncontrolled hypertension defined as systolic blood pressure (SBP) greater than 140 mmHg and/or diastolic blood pressure (DBP) greater than 90 mmHg; controlled hypertension defined as SBP lower than 140 mmHg and DBP lower than 90 mmHg but taking BP-lowering medicines; hypotension defined as SBP lower than 90 mmHg or DBP lower than 60 mmHg; and normotension, which comprised all other participants.

### Principal component analysis

Principal component analysis (PCA) was conducted on RNA-seq data normalised to transcripts per million (TPM), downloaded from the GTEx website in May 2025. Genes with TPM > 1 across more than 50% of whole-blood samples were considered expressed and included in the PCA model. The first 3 principal components (PCs) were extracted and evaluated for sex-by-hypertension interactions using a linear regression model adjusted for age, sex, body mass index (BMI), hypertension, diabetes diagnosis, smoking, and alcohol consumption, all included as covariates as follows:

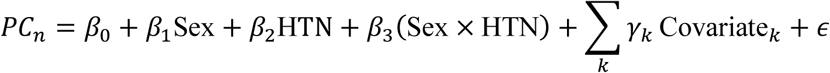

Where:

- *PC*_n_ is the responder, the n^th^ principal component.
- *β*_0_ is the intercept.
- Sex is coded as female = 1 and male = 0. HTN is coded as hypertension = 1 and normotension = 0.
- Covariates are age, body mass index (BMI), diabetes diagnosis, smoking, and alcohol consumption.
- Sex × HTN is the product of the two variables, representing the sex-by-hypertension interaction.
- *ε* is the error term.

Therefore, the values of PCs could be modelled in the following four circumstances after being adjusted for covariates:

1. **Normotensive male (**Sex = 0, HTN = 0): *β*_0_
2. **Normotensive female (**Sex = 1, HTN = 0): *β*_0_ + *β*_1_
3. **Hypertensive male (**Sex = 0, HTN = 1): *β*_0_ + *β*_2_
4. **Hypertensive female (**Sex = 1,HTN = 1): *β*_0_ + *β*_1_ + *β*_2_ + *β*_3_

Collectively, *β*_1_ is the simple effect of being female for non-hypertensive individuals, and *β*_2_ is the simple effect of hypertension for males. By contrast, *β*_3_ means the difference between the effect of hypertension in females versus the effect of hypertension in males.

For PCs with significant sex-by-hypertension interaction, the top 500 genes by variable contribution (loading) were extracted and analysed for pathway enrichment using Enrichr.^19^ The expression of these genes in different cell types detectable in whole blood was examined using the R package *celldex* (1.14.0).^20^

To simplify the model, two five-fold elastic net regression models were constructed using the genes commonly expressed in both the FHS and GTEx cohorts within the top 500 and top 200 (as a sensitivity analysis) contributing genes. We evaluated model performance by the correlation between observed and predicted PC values (the female hypertension co-expression signature) and by trends in mean squared error (MSE) versus regularisation strength (−log λ). We then tested sex-by-hypertension interactions in the GTEx cohort using the same linear regression model applied to the original observed PC values, with the female hypertension co-expression signature as the outcome.

We then extended the analysis to the FHS cohort to assess the model’s generalisability. The same female hypertension co-expression signatures derived from GTEx were calculated in the FHS cohort using coefficients derived from elastic net regression models trained on GTEx data as 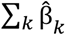 Gene*_k_*. Interactions between sex and both uncontrolled and controlled hypertension were assessed in the FHS cohort using the same linear regression framework adjusted by age, sex, BMI, diabetes diagnosis, smoking, and alcohol consumption, with normotension serving as the reference group.

### Identification of key regulatory transcription factors

TFs regulate gene expression by binding to short, conserved DNA sequences known as TF-binding motifs on promoter regions.^21^ To identify potential upstream regulators of the top-contributing genes for PCs, the promoter regions of these genes, defined as 2 kilobases upstream and 1 kilobase (kb) downstream (-2kb to +1kb) of the transcription start site (TSS), were extracted. TF-binding motif searching was performed using the findMotifs.pl function of Homer2 (4.11)^22^ on these promoter regions. We used results from both known-binding-motif matching and *de novo* binding-motif discovery to identify candidate upstream regulatory TFs.

We then downloaded all TF family members with significantly enriched binding motifs from the Human Transcription Factors database.^23^ Sex-by-hypertension interactions on the expression levels of the expressed members in whole blood were assessed in the GTEx cohort, with age, sex, BMI, hypertension, diabetes diagnosis, smoking, and alcohol consumption as covariates. TFs with significant sex-by-hypertension interactions were then used to construct an elastic net regression model, with the PC as the outcome and the expression levels of the selected TFs as predictors. To quantify the extent to which the expression of genes contributing to the PC could be explained by the included TFs, we evaluated the correlation between the TF-predicted PC values (namely, the TF expression score) and original observed PC values, as well as the proportion of variance explained by the predicted values. The significance of sex-by-hypertension interactions on the TF expression score was tested in both GTEx and FHS cohorts.

Finally, to examine the potential role of the identified TF family in BP, we constructed linear regression models of SBP versus TF expression scores, stratified by sex. To assess the potential confounding effects, we also constructed linear regression models using the adjusted SBP defined as the residual from a linear regression model with SBP as the response and age, BMI, diabetes status, smoking, BP-lowering medicines intake, and alcohol consumption as covariates.

### Co-expression analysis of artery, kidney, and heart

The associations between calculated female hypertension co-expression scores in the whole blood and the gene expression in 3 tissues with established roles in cardiovascular disease pathogenesis, the left ventricle of the heart, the coronary artery, and the kidney, were examined among male and female GTEx participants separately. Lowly expressed genes, defined as those with fewer than 10 read counts in more than half of the included samples, were excluded. Associations between predicted blood PC values and tissue-specific gene expression were assessed using DESeq2 (1.42.1)^24^ with coefficient size shrinkage implemented through the ASHR method.^25^ Genes with a false discovery rate (FDR) < 0.05 were considered significantly associated and subsequently subjected to pathway enrichment analysis using Enrichr.^19^ Gene Set Enrichment Analysis (GSEA) was conducted by the R package *ClusterProfiler* (4.12.6) with the Gene Ontology – Biological Processes (GO:BP) database.^26^

### Statistical analysis

Analyses of GTEx were performed in R 4.4, and analyses of FHS were performed in R 4.5 on the secure analytic platform constructed by Monash University. Five-fold elastic net regression models were trained using glmnet 5.0 function in R. Linear regression analyses were conducted using the lm function in R base. *: P < 0.05; **, P < 0.01; ***, P < 0.001.

## Results

### Gene set associated with hypertension in over 55-year-old females in two independent cohorts

To identify transcriptome signatures associated with hypertension in postmenopausal females (Figure 1A), we first extracted whole-blood transcriptome data from the human systemic transcriptome database GTEx.^14^ We restricted our sample to 193 participants (female n=61) older than 55 years without pre-existing CVD, representing an ageing population of predominantly postmenopausal women^15,16^ (Table 1, Figure 1A). Subsequently, a principal component analysis (PCA) was conducted to identify co-expression gene signatures from the whole-blood transcriptome. The first three principal components (PC1-3), which explained more than 50% of the total variance (Figure S1A), were extracted. We then constructed linear regression models with a sex-by-hypertension interaction term, using extracted PC values as response variables, to test for sex bias in these 3 PCs in the hypertension context, with common CVD risk factors (age, body mass index (BMI), smoking, alcohol consumption, and diabetes diagnosis) adjusted. Notably, PC1, but not PC2 or PC3, had a significant negative sex-by-hypertension interaction term (Figure 1B), suggesting that the underrepresentation of genes contributing to PC1 is a signature of hypertension in females but not in males.

Consistently with this, the top 500 genes by contribution (weight) to PC1 were uniformly positively correlated with PC1 (Table S1). The pathway enrichment analysis showed that these genes are involved in DNA repair, post-transcriptional modification, and insulin signalling (Figures 1C-D). Notably, these top PC1 contributing genes are highly expressed in B cells and CD4^+^ and CD8^+^ T cells, which are well-recognised blood pressure (BP) regulators (Figure S1B).^27^

**Figure 1.**
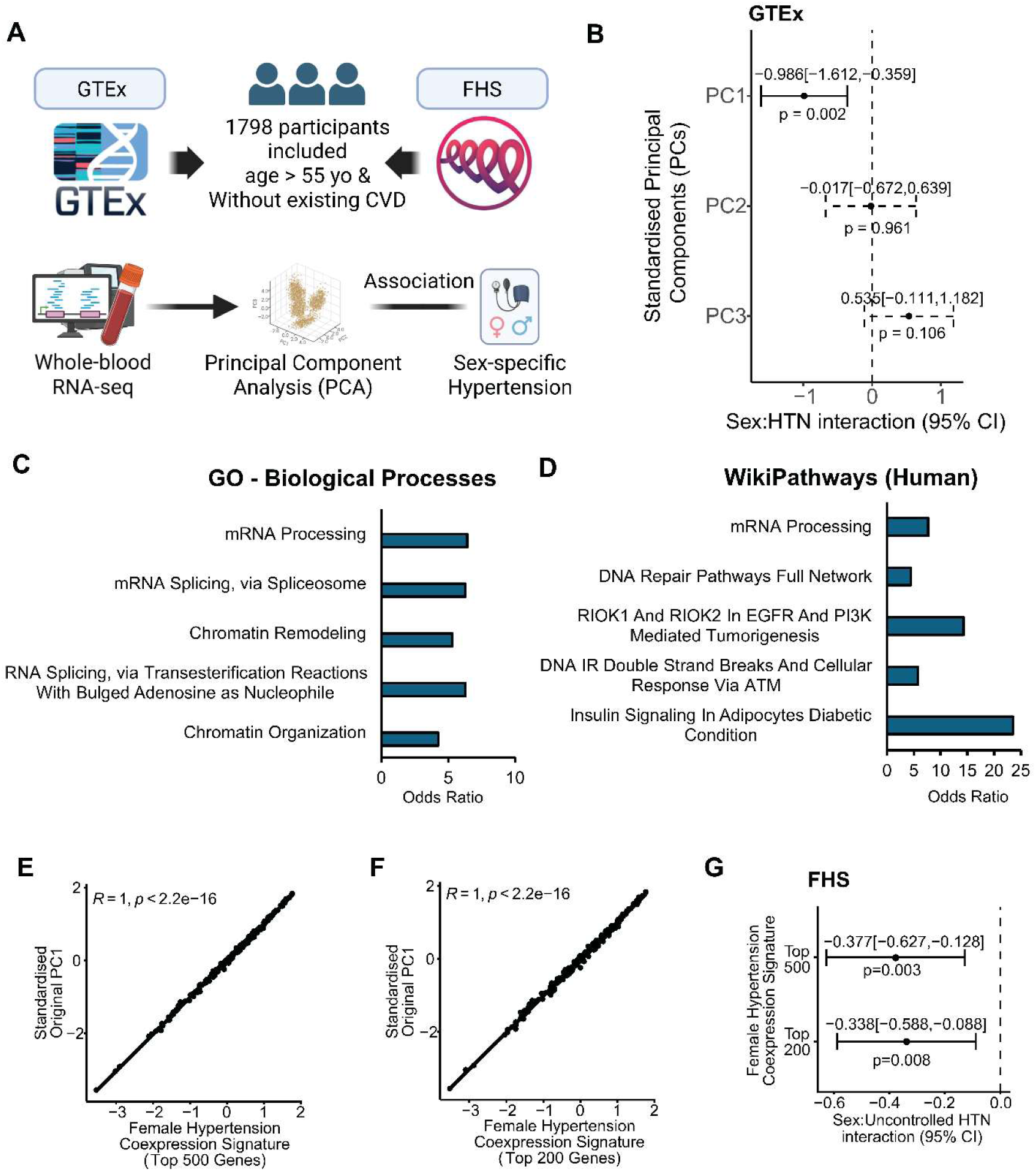
Sex-by-hypertension interaction on principal components (PCs) of transcriptome data. **A.** A summary of the study design. A total of 1,798 Participants, 193 from the Genotype-Tissue Expression (GTEx) discovery cohort and 1,605 from the Framingham Heart Study (FHS) validation cohort, aged> 55 years and without existing cardiovascular disease (CVD), were included in this study. **B.** Forest plot showing the sex-by-hypertension interactions on the standardised first 3 principal components (PC1-3) of GTEx whole blood transcriptome. The coefficients were presented as mean[lower 95% confidence limit, upper 95% confidence limit]. Models were adjusted for age, body mass index (BMI), smoking, alcohol consumption, and diabetes diagnosis. **C-D.** Pathway enrichment analysis of the top 500 PC1-contributing genes with the largest loadings using **C.** Gene Ontology (GO) – Biological Process and **D.** Wikipathways Database. **E-F.** The correlation between the original PC1 scores and the PC1 predictions made by **E.** top 500 and **F.** top 200 PC1-contributing genes using elastic net regression models, namely, the “female hypertension co-expression signature”. **G.** Forest plot showing the interactions between sex and uncontrolled hypertension on the calculated female hypertension co-expression signature scores based on top 500 and top 200 PC1-contributing genes from the whole blood transcriptome of the FHS cohort. The coefficients were presented as mean[lower 95% confidence limit, upper 95% confidence limit]. Models were adjusted for age, BMI, smoking, alcohol consumption, and diabetes diagnosis. n=193 for GTEx, n=1,605 for FHS.

**Table 1.** Demographics of included participants of the GTEx and FHS cohorts.

| GTEx |  |  |  |  |
| --- | --- | --- | --- | --- |
| Variable | Overall<br>N = 193 | Male<br>N = 132 | Female<br>N = 61 | p-value<br>(M vs F) |
| Age | 62 [59, 66] | 62 [59, 66] | 64 [60, 67] | 0.2 |
| BMI | 27.506 ± 4.003 | 27.962 ± 3.907 | 26.519 ± 4.062 | 0.023 |
| Hypertension | 121 (62.7%) | 78 (59.1%) | 43 (70.5%) | 0.13 |
| Diabetes | 50 (25.9%) | 34 (25.8%) | 16 (26.2%) | >0.9 |
| Alcohol consumption | 155 (80.3%) | 112 (84.8%) | 43 (70.5%) | 0.020 |
| Smoking | 123 (63.7%) | 89 (67.4%) | 34 (55.7%) | 0.12 |
| FHS |  |  |  |  |
| Variable | Overall<br>N = 1,605 | Male<br>N = 678 | Female<br>N = 927 | p-value<br>(M vs F) |
| Age | 66 [61, 73] | 66 [61, 72] | 66 [61, 73] | >0.9 |
| BMI | 28.165 ± 5.255 | 28.753 ± 4.595 | 27.735 ± 5.654 | <0.001 |
| Hypertension | 896 (55.8%) | 410 (60.4%) | 486 (52.4%) | 0.002 |
| Controlled HTN | 455 (28.3%) | 207 (30.5%) | 248 (26.8%) | 0.10 |
| Uncontrolled HTN | 441 (27.5%) | 203 (29.9%) | 238 (25.7%) | 0.059 |
| Diabetes | 196 (12.2%) | 109 (16.1%) | 87 (9.4%) | <0.001 |
| Alcohol consumption | 1,160 (72.3%) | 531 (78.3%) | 629 (67.9%) | <0.001 |
| Smoking | 114 (7.1%) | 44 (6.5%) | 70 (7.6%) | 0.4 |

Age is presented as Median [Q1, Q3]. BMI (body mass index) is presented as Mean ± SD. Categorical variables are presented as N (percentage). P-values of male versus female comparisons were calculated using the Wilcoxon-Mann-Whitney (WMW) test for continuous variables and Pearson's Chi-squared test for categorical variables. HTN, hypertension; F, female; M, male.

Subsequently, we examined the generalisability of the association between PC1 and the sex difference under hypertensive circumstances using another independent cohort study, the FHS.^18^ We extracted data from 1,640 participants aged over 55 years without a history of cardiovascular disease from the FHS Generation II Exam 8 datasets, with complete information collected from the clinical interview and examination. Because the FHS recorded menopausal status in the clinical interview, we reviewed these data and found that only 17 included female participants were premenopausal at the time of examination. This confirmed that the included participants aged over 55 years were postmenopausal females and age-matched males. After excluding these 17 premenopausal female participants as well as 18 participants with incomplete BP, alcohol consumption, smoking status, diabetes diagnosis data, or discontinued BP-lowering medicine intake, a total of 1,605 participants (female n=927) were retained for the subsequent analyses.

Due to the mathematical nature of PCA, PC1 is a weighted sum across all expressed genes, although most genes contribute negligibly. Moreover, the original PC1 is difficult to apply to other cohorts, such as FHS, that used a different platform and gene set. Therefore, to obtain a simpler and more biologically interpretable model, we constructed a reduced model by retaining the top 500 genes with the largest contributions to PC1 (Table S1) for subsequent analyses. We also conducted a sensitivity analysis using the top 200 contributing genes. We then used these reduced gene sets as predictors in two 5-fold elastic net regression models, which reweighted the genes according to the elastic net coefficients to predict PC1 values. As expected, given that these genes dominate the PC1 loadings, both models performed well and closely preserved the information in observed PC1 (Figures 1E-F, S1C-D; Table S2). Moreover, in analogous sex-by-hypertension interaction models adjusted for age, BMI, smoking, alcohol consumption, and diabetes diagnosis, both PC1 scores predicted (referred to as “female hypertension co-expression signatures”) from the top 500 and top 200 PC1-contributing genes showed significant negative sex-by-hypertension interactions in GTEx (Figure S1E). As a sensitivity analysis, we additionally included the first three genomic PCs as covariates to account for underlying population stratification, and the sex-by-hypertension interaction terms of female hypertension co-expression signature scores remained significantly negative (Table S3).

We then applied constructed elastic net regression models to another independent cohort, FHS, to calculate female hypertension co-expression signature scores. In the FHS cohort, detailed BP measurements and information on BP-lowering medication use enabled further classification of participants into four BP categories: uncontrolled hypertension, controlled hypertension, normotensive, and hypotension. Interestingly, in the FHS cohort, the interaction term between sex and uncontrolled hypertension was significantly negative on calculated female hypertension co-expression signature scores derived from both top-500- and top-200-based elastic net models, with the same covariates used in the GTEx analyses adjusted (Figure 1G). By contrast, the interaction term between sex and controlled hypertension was non-significant (Figure S1F). These findings suggest that this sex-dependent hypertension co-expression signature was primarily dependent on the presence of elevated BP, independent of medication. Overall, the results suggest that the underrepresentation of these top PC1-contributing genes is generalisable as a signature of female hypertension; thus, we termed these genes “female hypertension co-expression signature genes”.

### ETS family TFs are key regulators of the female-specific hypertensive transcriptome

Notably, 444 of the top 500 female hypertension co-expression signature genes were clustered together in a protein–protein interaction network analysis (Figure S1G, Table S4), indicating that these genes are highly functionally related. This observation led us to hypothesise that a common set of transcriptional regulators could underlie their coordinated expression patterns. To identify potential upstream regulators, we extracted the promoter regions of the top female hypertension co-expression signature genes and searched for enriched conserved transcription factor (TF) DNA-binding motifs^21^ in these regions. The results of the *de novo* search showed that the motifs of E26 transformation-specific (ETS) family TFs were highly enriched in promoter regions of both the top 500 and top 200 female hypertension co-expression signature genes (Figures 2A-B). Previous reports have demonstrated their critical role in defending against external perturbations such as lipopolysaccharide (LPS) and mitigating induced injuries.^28-30^ Consistently, results from known motif database matching also showed that the DNA-binding motifs of ETS family TFs were the most significantly enriched (Figures S2A-B).

**Figure 2.**
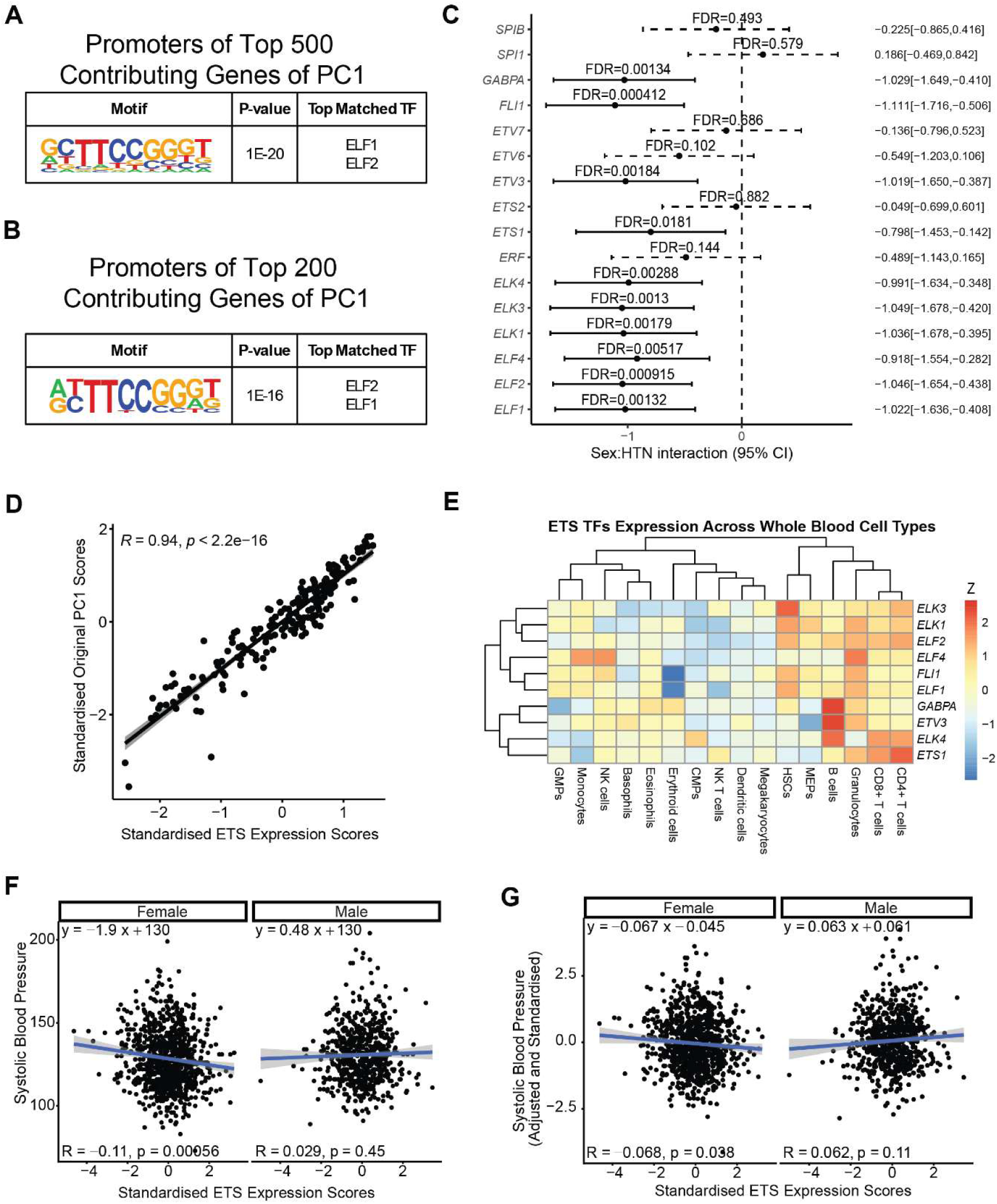
Sex-by-hypertension interaction on expression of ETS family transcription factors (TFs). **A-B.** Results of *de novo* DNA-binding motif searching on the promoter regions defined as 2kb upstream to 1kb downstream regions from the transcription start site (-2 kb to +1 kb from TSS) of **A.** top 500 and **B.** top 200 PC1-contributing genes. **C.** Forest plot showing the sex-by-hypertension interactions on the expression levels of 16 ETS family TFs expressed in whole blood. The coefficients were presented as mean[lower 95% confidence limit, upper 95% confidence limit]. P-values shown on the plot were adjusted by the Benjamini-Hochberg procedure. Models were adjusted for age, body mass index (BMI), smoking, alcohol consumption, and diabetes diagnosis. **D.** The correlation between the original PC1 scores and the PC1 predictions made by 10 ETS family TFs, namely, the “ETS expression score” with significant negative sex-by-hypertension interactions. **E**. Expression of the 10 ETS family TFs by cell type in whole blood. CMPs, common myeloid progenitors; GMPs, granulocyte-macrophage progenitors; HSCs, hematopoietic stem cells; MEPs, megakaryocyte-erythroid progenitors. **F-G.** Correlation between ETS expression scores and **F**. unadjusted and **G.** systolic blood pressure (SBP) adjusted by age, BMI, smoking, alcohol consumption, diabetes diagnosis, and BP-lowering medication intake, in included male and female participants of FHS. n=193 for GTEx, n=1,605 for FHS.

Subsequently, we extracted all 29 ETS-family TFs from the Human Transcription Factors database^23^ to assess their expression levels in whole blood from the GTEx cohort. A set of linear models, including a sex-by-hypertension interaction term and adjustment for age, BMI, alcohol consumption, smoking, and diabetes diagnosis, was constructed using the expression levels of these genes as the response. Sixteen out of all twenty-nine (55%) ETS members were expressed in whole blood, and ten members had a significant negative sex-by-hypertension interaction term (Figure 2C). These results are consistent with the enrichment of ETS family DNA-binding motifs in the promoter regions of the top female hypertension co-expression signature genes.

To further assess the connection between ETS family TFs and the top female hypertension co-expression signature genes, we constructed an elastic net regression model to test whether expression of these 10 sex-by-hypertension-interacting ETS TFs could predict PC1 values. This approach hypothesises that if ETS TFs are upstream regulators of these top PC1-contributing female hypertension co-expression signature genes, their expression profiles should capture the coordinated variation represented by PC1. Remarkably, the PC1 predictions based on these 10 ETS TFs (referred to as “ETS expression scores”) were highly accurate, as they captured around 88% (R = 0.94, Figures 2D and S2C; Table S5) of the original observed PC1’s variance. In the GTEx cohort, the sex-by-hypertension interaction on the ETS expression scores was also significantly negative (β=-1.080[-1.656,-0.504]; *p=3.16E-4*; Table S6). Moreover, like the top 500 and top 200 female hypertension co-expression signature genes, these ETS family TFs were also highly expressed in CD4^+^ and CD8^+^ T cells and B cells (Figure 2E). Collectively, these findings suggest that the identified female hypertension co-expression genes and the ETS family TFs likely form a transcriptional program primarily active in B cells and T cells, which might be suppressed in the development of hypertension in females.

We then extended our findings to the FHS cohort. We calculated ETS expression scores in the FHS cohort using the elastic net regression coefficients estimated from the ETS TF-based elastic net regression model constructed in GTEx. The FHS cohort has more detailed BP-based stratification, allowing us to explore BP control instead of hypertensive status. Indeed, the interaction term between sex and uncontrolled hypertension for ETS expression scores was significantly negative (β=-0.247 [-0.495,-0.001]; *p*=4.97E-2; Table S7), while the interaction term between sex and controlled hypertension for ETS expression scores was non-significant (β=-0.119 [-0.307,0.069]; *p*=2.15E-1; Table S7). Therefore, we stratified participants by sex and examined the correlation between ETS expression scores and SBP separately in males and females. Remarkably, ETS expression scores were only significantly associated with lower SBP in female, but not male, FHS participants (Figure 2F). In an analogous model using SBP adjusted for age, BMI, alcohol consumption, smoking status, diabetes diagnosis, and BP-lowering medication in the FHS cohort, the ETS expression scores were also significantly negatively correlated with the adjusted SBP in females, but not in males (Figure 2G). These results showed that the association between ETS family expression and lower SBP observed in female participants is independent of BP-lowering medication.

### The female-specific hypertensive transcriptome is associated with pro-inflammatory and pro-fibrotic transcriptomes in kidneys and artery

Finally, we assessed whether blood-derived female hypertension co-expression signature scores representing genes underrepresented in hypertensive females over 55 were associated with tissue-specific transcriptional alterations relevant to cardiovascular disease development. We analysed available transcriptomic data from included female and male GTEx participants in the left ventricle, coronary artery, and renal cortex, which are closely linked to cardiovascular diseases. (n=123 for heart; 66 for coronary artery, and 33 for kidney cortex). Genes with false-discovery rate (FDR) < 0.05 and log_2_-transformed fold change (log_2_FC) > 0 were considered as genes significantly overrepresented by female hypertension signature scores, and genes with log_2_FC<0 and FDR<0.05 as significantly underrepresented.

Unexpectedly, we found that no gene significantly correlated with female hypertension co-expression signature scores in the left ventricle in either males or females (Figure 3A). However, in the coronary artery, we found 3 genes significantly overrepresented and 18 genes underrepresented by higher female hypertension co-expression signature scores trained by the top 500 genes (Table S8, Figure 3A) in females. The 18 significantly underrepresented genes in the coronary artery are mainly pro-inflammatory, such as *CCL3* and *CCL19* (Figure 3B). By contrast, we identified no significantly overrepresented or underrepresented genes with hypertension co-expression signature scores in the male coronary artery. These findings were consistent with gene set enrichment analysis (GSEA), in which the Gene Ontology (GO) term “adaptive immune response” was significantly enriched only among genes that were underrepresented with higher female hypertension co-expression signature scores in coronary artery tissue of females (Figure 3C), but not males (Figure 3D). In the kidney cortex of females, 380 genes were significantly underrepresented, and 561 were significantly overrepresented with higher female hypertension co-expression signature scores (Table S8, Figure 3A). Genes that were underrepresented at higher female hypertension co-expression signature scores were enriched for profibrotic pathways (Figure 3E). Conversely, underrepresented genes at higher female hypertension co-expression signature scores were primarily involved in small-molecule transport (Figure 3F), a hallmark of preserved renal function.^31,32^ By comparison, in the kidney cortex of males, only two genes were overrepresented, and three genes were underrepresented in association with higher co-expression signature scores (Table S8). Consistently, the GO term “collagen fibril organization” including genes like *COL1A1,* was significantly enriched only among genes that were underrepresented with higher female hypertension co-expression signature scores in the kidney of females (Figure 3G), but not males (Figure 3H). The GO term “inorganic anion transport” was significantly enriched only among genes that were overrepresented with higher female hypertension co-expression signature scores in females, but with no significant enrichment observed in males (Figure S3).

**Figure 3.**
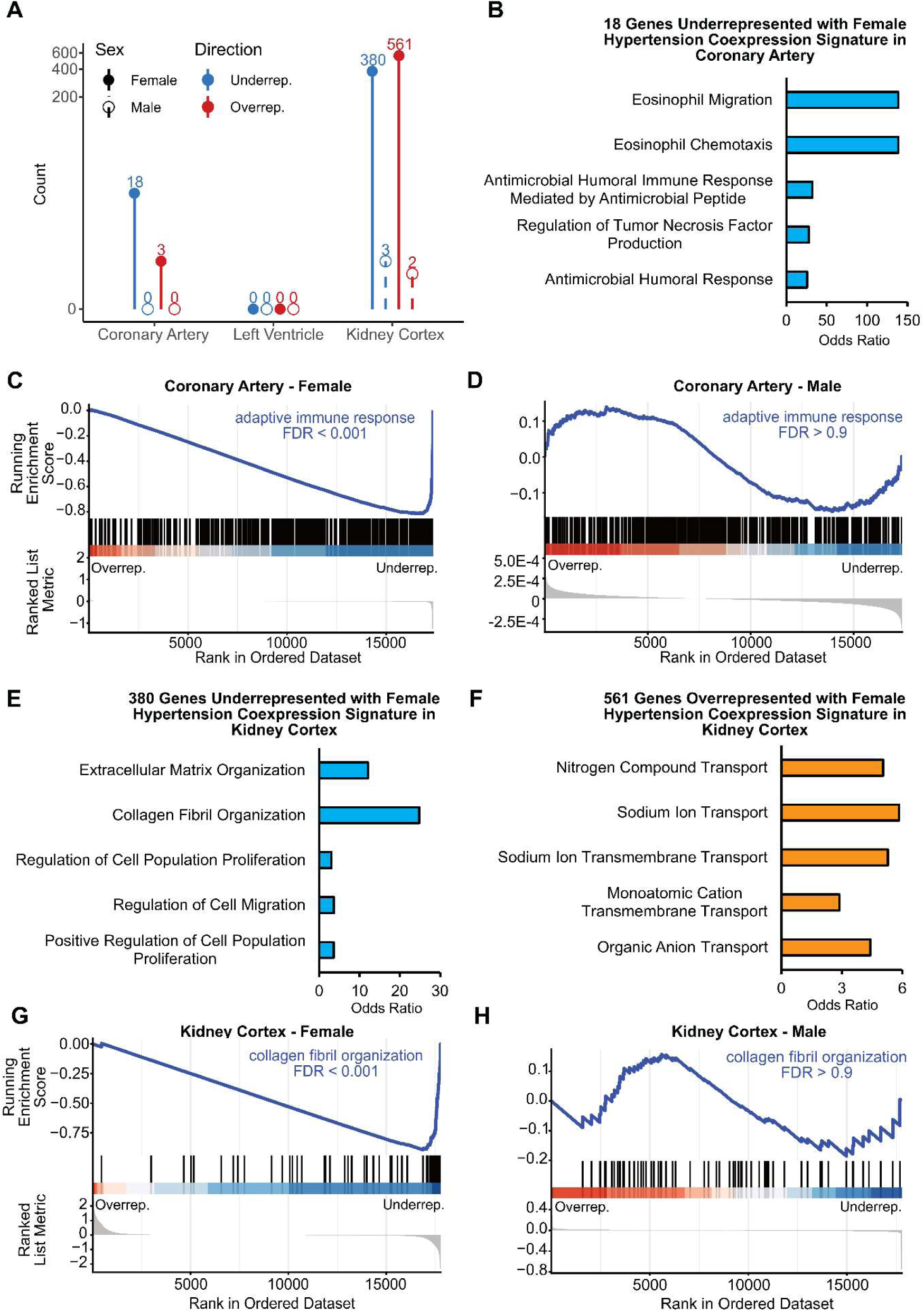
Association between the female hypertension co-expression signature and systematic transcriptomic alterations. **A.** A summary of the number of genes over- and under-represented with higher female hypertension co-expression signature scores in males (dashed lines) and females (solid lines). **B.** Pathway enrichment analysis of the genes underrepresented with higher female hypertension co-expression signature scores in female coronary artery. **C-D.** Gene Set Enrichment Analysis (GSEA) of the Gene Ontology (GO) term “adaptive immune response” in the coronary artery transcriptome from **C.** females and **D.** males. The term was enriched in genes underrepresented with higher female hypertension co-expression signature scores in females, but not in males. **E-F.** Pathway enrichment analysis of the genes **E.** underrepresented, and **F.** overrepresented with higher female hypertension co-expression signature scores in the female kidney cortex. **G-H.** GSEA of the GO term “collagen fibril organization” in the kidney cortex transcriptome from **C.** females and **D.** males. n=123 for heart; 66 for coronary artery; and 33 for kidney cortex.

## Discussion

In this study, we identified a blood transcriptomic signature that is consistently under-expressed in postmenopausal females with hypertension – but not in age-matched males – across two independent cohorts with> 1,600 participants. This signature is driven by a coordinated set of genes regulated by ETS family transcription factors: ten ETS TFs alone captured most of its variation and were significantly underrepresented in hypertensive females in both cohorts. Both the signature co-expression genes and ETS regulators are highly expressed in B cells and CD4^+^ and CD8^+^ T cells, immune populations with established roles in BP regulation.^33^ Collectively, these findings point to a female-specific, ETS-regulated transcriptional programme in circulating immune cells whose deficiency is associated with hypertension in postmenopausal women.

Notably, identified female hypertension co-expression genes were enriched for metabolic homeostasis, post-transcriptional regulation, and DNA repair. Dysregulation of DNA repair^34,35^ contributes to reproductive ageing in females, reducing estrogen production, which has anti-inflammatory and anti-fibrotic effects and can thus lower cardiovascular risk.^36^ Indeed, the included female participants older than 55 years old in GTEx represent a predominant post-menopausal female population.^15,16,37^ Furthermore, analysis of the available menopause status data from the FHS confirmed that only approximately 1-2% of female participants older than 55 years were still premenopausal, which were excluded from subsequent analyses in FHS to avoid potential confounding by later menopause. Thus, these findings collectively indicated that the observed association between the underrepresentation of this signature and hypertension in older females likely reflects the gradual loss of a cardioprotective mechanism following menopause.

Our analyses revealed that the identified female hypertension co-expression signature was associated with lower expression of pro-inflammatory genes in the artery and pro-fibrotic genes in the kidney in females but not males, suggesting preserved artery and kidney function, which are essential for cardiovascular health. A study showed that older hypertensive females have significantly compromised kidney function compared to age-matched hypertensive males^38^, suggesting that declining renal function may contribute to the female-specific hypertension context. Collectively, these observations indicate that the underrepresentation of the female hypertension co-expression signature is associated with the development of hypertension in postmenopausal females.

ETS family TFs play a vital role in defence responses against external stressors, such as viruses and LPS, and can thus suppress further tissue damage.^29,30,39^ Particularly, detrimental stimuli such as LPS have been implicated in the development of hypertension.^40^ Additionally, the ETS family member GABPA, known to help reduce oxidative stress,^41^ was significantly underrepresented in heart failure conditions.^42^ Therefore, the underrepresentation of ETS family TFs in older hypertensive females may indicate a diminished capacity to respond to and eliminate detrimental external perturbations to the cardiovascular system. Moreover, in our study, in both GTEx and FHS cohorts, the underrepresentation of ETS-based PC1 prediction, named “ETS expression scores”, was associated with hypertension in females. In FHS participants with SBP data available, ETS expression scores were also negatively correlated with systolic blood pressure in females, but not males, and this association was independent of age, BMI, diabetes, alcohol consumption, smoking, or BP-lowering medication. These results suggest that ETS family TF expression in blood plays a role in BP regulation in females, independent of BP-lowering medication intake.

We also acknowledge several limitations of this study. Firstly, all participants from the Generation II Exam8 of the FHS cohort consisted exclusively of individuals of European ancestry. While GTEx participants were from different ancestral backgrounds, they were still predominantly of European ancestry. Therefore, future studies involving more ancestrally diverse populations with a higher presence of non-European participants are warranted to establish the broader applicability of this female-specific hypertension signature. Secondly, our study didn’t establish causal relationships between female hypertension, PC1-contributing genes, and ETS family TFs. Therefore, further studies using animal or *ex vivo* human systems are needed to confirm whether perturbation on this transcriptional network contributes directly to the development of female hypertension and its associated cardiovascular phenotypes. Thirdly, because the GTEx did not provide detailed BP data, we cannot further subgroup the hypertension in this dataset. Finally, because neither cohort provided blood cell count data alongside the whole-blood transcriptomic data, we were unable to determine whether the identified female hypertension-associated co-expression signature was related to alterations in blood cell composition, particularly in CD4+ and CD8+ T cells, which have been reported in hypertension.^43,44^ Nevertheless, sex differences in these and other cell populations have not been described. Therefore, although we cannot completely exclude the potential influence of blood cell composition, the observed co-expression signature is likely to be driven primarily by shifts in transcriptional reprogramming rather than shifts in circulating immune cell numbers.

Overall, we identified a transcriptional signature consistently underrepresented in hypertensive females in two independent cohorts, with genes involved in metabolic homeostasis, post-transcriptional regulation, and DNA repair significantly enriched. The reproducibility of this signature across independent cohorts highlights dysfunction in the identified female hypertensive co-expressed genes and their upstream ETS-family TFs in the pathogenesis of female hypertension. Lower expression of these genes was also associated with pro-inflammatory, pro-fibrotic transcriptomes in arterial and renal tissues, both central to cardiovascular disease pathogenesis. Beyond its biological significance, this regulatory network may also serve as a target for developing preventive and therapeutic approaches for female hypertension.

## Acknowledgements

This research used the Framingham Heart Study (FHS) and the Genotype-Tissue Expression (GTEx) resources under Application Number 142837-1 and 147531-2. The analytical plan for the FHS cohort was approved by Monash University Human Research Ethics Committee (MUHREC) under the Application Number 48482. Monash eResearch, including the M3 servers, supported this research.

## Author Contributions

Concept and design: CY, FZM. Acquisition, analysis and interpretation of data: CY, LCT, FZM. Drafting of manuscript: CY, FZM. Critical revision of manuscript: CY, JL, TP, LCT; Statistical analysis: CY. Analytical code development and optimisation: CY, JL, TP. Obtained funding, access to FHS and GTEx, and supervision: FZM. All authors read and approved the manuscript.

## Data availability

Data from the Framingham Heart Study (FHS) and the Genotype-Tissue Expression (GTEx) project are accessible via approved applications through the National Institutes of Health (NIH) dbGaP Authorised Access System. The R codes are available at https://github.com/ChrYang/FemaleHypertension.

## References

1. Brauer, M., Roth, G.A., Aravkin, A.Y., Zheng, P., Abate, K.H., Abate, Y.H., Abbafati, C., Abbasgholizadeh, R., Abbasi, M.A., Abbasian, M., et al. (2024). Global burden and strength of evidence for 88 risk factors in 204 countries and 811 subnational locations, 1990–2021: a systematic analysis for the Global Burden of Disease Study 2021. The Lancet 403, 2162–2203. 10.1016/s0140-6736(24)00933-4.

2. Ostchega, Y., Fryar, C.D., Nwankwo, T., and Nguyen, D.T. (2020). Hypertension Prevalence Among Adults Aged 18 and Over: United States, 2017-2018. NCHS Data Brief, 1–8.

3. Osude, N., Durazo-Arvizu, R., Markossian, T., Liu, K., Michos, E.D., Rakotz, M., Wozniak, G., Egan, B., and Kramer, H. (2021). Age and sex disparities in hypertension control: The multi-ethnic study of atherosclerosis (MESA). Am J Prev Cardiol 8, 100230. 10.1016/j.ajpc.2021.100230.

4. Zanchetti, A., Facchetti, R., Cesana, G.C., Modena, M.G., Pirrelli, A., and Sega, R. (2005). Menopause-related blood pressure increase and its relationship to age and body mass index: the SIMONA epidemiological study. J Hypertens 23, 2269–2276. 10.1097/01.hjh.0000194118.35098.43.

5. Ryczkowska, K., Adach, W., Janikowski, K., Banach, M., and Bielecka-Dabrowa, A. (2023). Menopause and women’s cardiovascular health: is it really an obvious relationship? Arch Med Sci 19, 458–466. 10.5114/aoms/157308.

6. El Khoudary, S.R., Aggarwal, B., Beckie, T.M., Hodis, H.N., Johnson, A.E., Langer, R.D., Limacher, M.C., Manson, J.E., Stefanick, M.L., and Allison, M.A. (2020). Menopause Transition and Cardiovascular Disease Risk: Implications for Timing of Early Prevention: A Scientific Statement From the American Heart Association. Circulation 142. 10.1161/cir.0000000000000912.

7. Yanes, L.L., and Reckelhoff, J.F. (2011). Postmenopausal hypertension. Am J Hypertens 24, 740–749. 10.1038/ajh.2011.71.

8. Ji, H., Kim, A., Ebinger, J.E., Niiranen, T.J., Claggett, B.L., Bairey Merz, C.N., and Cheng, S. (2020). Sex Differences in Blood Pressure Trajectories Over the Life Course. JAMA Cardiol 5, 19–26. 10.1001/jamacardio.2019.5306.

9. Ji, H., Niiranen, T.J., Rader, F., Henglin, M., Kim, A., Ebinger, J.E., Claggett, B., Merz, C.N.B., and Cheng, S. (2021). Sex Differences in Blood Pressure Associations With Cardiovascular Outcomes. Circulation 143, 761–763. 10.1161/circulationaha.120.049360.

10. Matetic, A., Shamkhani, W., Rashid, M., Volgman, A.S., Van Spall, H.G.C., Coutinho, T., Mehta, L.S., Sharma, G., Parwani, P., Mohamed, M.O., and Mamas, M.A. (2021). Trends of Sex Differences in Clinical Outcomes After Myocardial Infarction in the United States. CJC Open 3, S19–s27. 10.1016/j.cjco.2021.06.012.

11. Guzik, T.J., Nosalski, R., Maffia, P., and Drummond, G.R. (2024). Immune and inflammatory mechanisms in hypertension. Nat Rev Cardiol 21, 396–416. 10.1038/s41569-023-00964-1.

12. Staessen, J.A., van der Heijden-Spek, J.J., Safar, M.E., Den Hond, E., Gasowski, J., Fagard, R.H., Wang, J.G., Boudier, H.A.S., and Van Bortel, L.M. (2001). Menopause and the characteristics of the large arteries in a population study. Journal of Human Hypertension 15, 511–518. 10.1038/sj.jhh.1001226.

13. Bruno, R.-M., Varbiro, S., Pucci, G., Nemcsik, J., Lønnebakken, M.T., Kublickiene, K., Schluchter, H., Park, C., Mozos, I., Guala, A., et al. (2023). Vascular function in hypertension: does gender dimension matter? Journal of Human Hypertension 37, 634–643. 10.1038/s41371-023-00826-w.

14. Lonsdale, J., Thomas, J., Salvatore, M., Phillips, R., Lo, E., Shad, S., Hasz, R., Walters, G., Garcia, F., Young, N., et al. (2013). The Genotype-Tissue Expression (GTEx) project. Nature Genetics 45, 580–585. 10.1038/ng.2653.

15. Charafi, L., Bolling, K., Schroader, B.K., and Halvorson, L. (2025). Characterization and Treatment Patterns of Peri/Menopausal and Postmenopausal Women with and Without Vasomotor Symptoms in a Retrospective Database Study. Womens Health Rep (New Rochelle) 6, 742–751. 10.1177/26884844251366113.

16. Monteiro Fernandes, A., Varino, F., F, S.S., Baronet, P., and Félix Almeida, F. (2025). Hospitalization in women with psychosis: An age-based proxy analysis for menopause. J Psychiatr Res 191, 262–264. 10.1016/j.jpsychires.2025.09.067.

17. McEvoy, J.W., McCarthy, C.P., Bruno, R.M., Brouwers, S., Canavan, M.D., Ceconi, C., Christodorescu, R.M., Daskalopoulou, S.S., Ferro, C.J., Gerdts, E., et al. (2024). 2024 ESC Guidelines for the management of elevated blood pressure and hypertension: Developed by the task force on the management of elevated blood pressure and hypertension of the European Society of Cardiology (ESC) and endorsed by the European Society of Endocrinology (ESE) and the European Stroke Organisation (ESO). European Heart Journal 45, 3912–4018. 10.1093/eurheartj/ehae178.

18. Mahmood, S.S., Levy, D., Vasan, R.S., and Wang, T.J. (2014). The Framingham Heart Study and the epidemiology of cardiovascular disease: a historical perspective. Lancet 383, 999–1008. 10.1016/s0140-6736(13)61752-3.

19. Kuleshov, M.V., Jones, M.R., Rouillard, A.D., Fernandez, N.F., Duan, Q., Wang, Z., Koplev, S., Jenkins, S.L., Jagodnik, K.M., Lachmann, A., et al. (2016). Enrichr: a comprehensive gene set enrichment analysis web server 2016 update. Nucleic Acids Research 44, W90–W97. 10.1093/nar/gkw377.

20. Aran, D., Looney, A.P., Liu, L., Wu, E., Fong, V., Hsu, A., Chak, S., Naikawadi, R.P., Wolters, P.J., Abate, A.R., et al. (2019). Reference-based analysis of lung single-cell sequencing reveals a transitional profibrotic macrophage. Nature Immunology 20, 163–172. 10.1038/s41590-018-0276-y.

21. Inukai, S., Kock, K.H., and Bulyk, M.L. (2017). Transcription factor-DNA binding: beyond binding site motifs. Curr Opin Genet Dev 43, 110–119. 10.1016/j.gde.2017.02.007.

22. Heinz, S., Benner, C., Spann, N., Bertolino, E., Lin, Y.C., Laslo, P., Cheng, J.X., Murre, C., Singh, H., and Glass, C.K. (2010). Simple Combinations of Lineage-Determining Transcription Factors Prime cis-Regulatory Elements Required for Macrophage and B Cell Identities. Molecular Cell 38, 576–589. 10.1016/j.molcel.2010.05.004.

23. Lambert, S.A., Jolma, A., Campitelli, L.F., Das, P.K., Yin, Y., Albu, M., Chen, X., Taipale, J., Hughes, T.R., and Weirauch, M.T. (2018). The Human Transcription Factors. Cell 172, 650–665. 10.1016/j.cell.2018.01.029.

24. Love, M.I., Huber, W., and Anders, S. (2014). Moderated estimation of fold change and dispersion for RNA-seq data with DESeq2. Genome Biology 15, 550. 10.1186/s13059-014-0550-8.

25. Stephens, M. (2017). False discovery rates: a new deal. Biostatistics 18, 275–294. 10.1093/biostatistics/kxw041.

26. Wu, T., Hu, E., Xu, S., Chen, M., Guo, P., Dai, Z., Feng, T., Zhou, L., Tang, W., Zhan, L., et al. (2021). clusterProfiler 4.0: A universal enrichment tool for interpreting omics data. The Innovation 2, 100141. 10.1016/j.xinn.2021.100141.

27. Dinakis, E., O’Donnell, J.A., and Marques, F.Z. (2024). The gut–immune axis during hypertension and cardiovascular diseases. Acta Physiologica 240, e14193. 10.1111/apha.14193.

28. Yuan, L., Nikolova-Krstevski, V., Zhan, Y., Kondo, M., Bhasin, M., Varghese, L., Yano, K., Carman, C.V., Aird, W.C., and Oettgen, P. (2009). Antiinflammatory effects of the ETS factor ERG in endothelial cells are mediated through transcriptional repression of the interleukin-8 gene. Circ Res 104, 1049–1057. 10.1161/circresaha.108.190751.

29. Seifert, L.L., Si, C., Saha, D., Sadic, M., de Vries, M., Ballentine, S., Briley, A., Wang, G., Valero-Jimenez, A.M., Mohamed, A., et al. (2019). The ETS transcription factor ELF1 regulates a broadly antiviral program distinct from the type I interferon response. PLoS Pathog 15, e1007634. 10.1371/journal.ppat.1007634.

30. Wang, D.-M., Yang, J., Ansari, M.O., Arbieva, Z., Maienschein-Cline, M., Natarajan, V., Malik, A.B., and Tiruppathi, C. (2026). CaMKK&#x3b2; regulates transcription factor Elf2 gene methylation to maintain endothelial junctional barrier integrity. iScience 29. 10.1016/j.isci.2025.114255.

31. Beckerman, P., Qiu, C., Park, J., Ledo, N., Ko, Y.A., Park, A.D., Han, S.Y., Choi, P., Palmer, M., and Susztak, K. (2017). Human Kidney Tubule-Specific Gene Expression Based Dissection of Chronic Kidney Disease Traits. EBioMedicine 24, 267–276. 10.1016/j.ebiom.2017.09.014.

32. Xie, J.X., Li, X., and Xie, Z. (2013). Regulation of renal function and structure by the signaling Na/K-ATPase. IUBMB Life 65, 991–998. 10.1002/iub.1229.

33. Guzik, T.J., Nosalski, R., Maffia, P., and Drummond, G.R. (2024). Immune and inflammatory mechanisms in hypertension. Nature Reviews Cardiology 21, 396–416. 10.1038/s41569-023-00964-1.

34. Turan, V., and Oktay, K. (2020). BRCA-related ATM-mediated DNA double-strand break repair and ovarian aging. Human Reproduction Update 26, 43–57. 10.1093/humupd/dmz043.

35. Oktay, K., Turan, V., Titus, S., Stobezki, R., and Liu, L. (2015). BRCA Mutations, DNA Repair Deficiency, and Ovarian Aging. Biol Reprod 93, 67. 10.1095/biolreprod.115.132290.

36. Xiang, D., Liu, Y., Zhou, S., Zhou, E., and Wang, Y. (2021). Protective Effects of Estrogen on Cardiovascular Disease Mediated by Oxidative Stress. Oxid Med Cell Longev 2021, 5523516. 10.1155/2021/5523516.

37. McKinlay, S.M. (1996). The normal menopause transition: an overview. Maturitas 23, 137–145. 10.1016/0378-5122(95)00985-X.

38. Beauregard, N., Vinson, A.J., McIsaac, D.I., Girard, C., Sood, M.M., Kendall, C.E., Sweet, A., Singla, R., Motazedian, P., Hundemer, G.L., et al. (2026). Older females with late-onset hypertension are at higher CKD risk than males. Clinical Kidney Journal 19. 10.1093/ckj/sfag071.

39. Luo, C.T., Osmanbeyoglu, H.U., Do, M.H., Bivona, M.R., Toure, A., Kang, D., Xie, Y., Leslie, C.S., and Li, M.O. (2017). Ets transcription factor GABP controls T cell homeostasis and immunity. Nature Communications 8, 1062. 10.1038/s41467-017-01020-6.

40. Grylls, A., Seidler, K., and Neil, J. (2021). Link between microbiota and hypertension: Focus on LPS/TLR4 pathway in endothelial dysfunction and vascular inflammation, and therapeutic implication of probiotics. Biomedicine & Pharmacotherapy 137, 111334. 10.1016/j.biopha.2021.111334.

41. Niopek, K., Üstünel, B.E., Seitz, S., Sakurai, M., Zota, A., Mattijssen, F., Wang, X., Sijmonsma, T., Feuchter, Y., Gail, A.M., et al. (2017). A Hepatic GAbp-AMPK Axis Links Inflammatory Signaling to Systemic Vascular Damage. Cell Reports 20, 1422–1434. 10.1016/j.celrep.2017.07.023.

42. He, P., Deng, L., and Wu, K. (2025). Multi-omics approach reveals CCND1, GABPA, HIF1A, and SOX6 as key regulators and prognostic markers in heart failure. Hereditas 162, 165. 10.1186/s41065-025-00536-y.

43. Alexander, M.R., Dale, B.L., Smart, C.D., Elijovich, F., Wogsland, C.E., Lima, S.M., Irish, J.M., and Madhur, M.S. (2023). Immune Profiling Reveals Decreases in Circulating Regulatory and Exhausted T Cells in Human Hypertension. JACC Basic Transl Sci 8, 319–336. 10.1016/j.jacbts.2022.09.007.

44. Dinakis, E., Rhys-Jones, D., Muir, J., O’Donnell, J.A., and Marques, F.Z. (2026). Deep immunophenotyping reveals associations with human blood pressure. medRxiv, 2026.2001.2014.26344062. 10.64898/2026.01.14.26344062.

